# Accelerometer-measured weekend catch-up sleep and incident dementia: a prospective cohort study

**DOI:** 10.1101/2025.08.27.25334608

**Authors:** Hui Chen, Ting Shen, Mengjia Zhao, Lusha Tong, Guiyu Lu, Wenchang Yang, Fengchang Li, Chengwu Feng, Geng Zong, Xiao Tan, Tianyi Huang, Changzheng Yuan

## Abstract

**Objective:** To investigate whether accelerometer-measured weekend catch-up sleep, defined as extending sleep on weekends to compensate for weekday sleep inadequacy, is associated with incident dementia.

**Design:** Prospective cohort study.

**Setting:** UK Biobank.

**Participants:** 83,776 dementia-free adults aged 50 years or older (mean age: 63.3 years, standard deviation: 6.7 years; 56.5% female) were included at the time of seven-day wrist-worn accelerometer assessment (2013-2015), followed until December 2022 for all-cause dementia incidence.

**Main outcome measures:** Daily sleep duration was derived from the seven-day accelerometer data using a machine learning approach. Weekend catch-up sleep was defined as the weekend-weekday difference in average sleep duration. Incident all-cause dementia was identified from linked healthcare data. Cox proportional hazard regression models was used to calculate hazard ratios (HRs) and 95% confidence intervals (CIs).

**Results:** During a total of 667,928 person-years (median follow-up: 8.0 years), 713 participants developed all-cause dementia. A significant lower risk of dementia was observed among individuals with a moderate level of weekend catch-up sleep (1-1.5 hours/day). Specifically, compared with none or ≤0.5 hour, the multi-variable adjusted HRs and 95% CIs across increasing categories of weekend catch-up sleep (>0.5-1, >1-1.5, >1.5-2, and >2 hours) were 0.91 (0.74-1.11), 0.64 (0.49-0.86), 0.84 (0.60-1.16), and 0.83 (0.60-1.16), respectively. The association was stronger among participants with a weekday sleep duration <8 hours (HR: 0.49, 0.29-0.81 for >1-1.5 hours), but non-significant in those with weekday sleep durations ≥8 hours (HR: 0.72, 0.51-1.01 for >1-1.5 hours, *P*-interaction = 0.039).

**Conclusions:** Moderate accelerometer-tracked weekend catch-up sleep was significantly associated with lower risk of all-cause dementia, particularly among individuals with less weekday sleep. Future studies are needed to confirm these findings and identify the optimal sleep compensation strategies for dementia prevention.

## Introduction

Dementia is a leading cause of disability among older adults and poses increasing healthcare burdens worldwide, with over 50 million cases in 2019.^1–3^ Given that there is currently limited cure to reverse its progression, identifying modifiable lifestyle factors is crucial for effective prevention of dementia.^1,4^ Healthy sleep duration emerged as a key component of healthy lifestyle in maintaining brain health, and both short and long sleep durations have been linked to cognitive decline and a higher risk of dementia.^5^

In individuals with insufficient sleep during the week, “weekend catch-up sleep”—the practice of extending sleep on weekends—has become a way to compensate for sleep deficits accumulated from weekday demands.^6^ While prior research has linked weekend catch-up sleep to various metabolic health status,^7–9^ its long-term effects on brain health are largely unexplored. A cross-sectional study of 215 older adults reported an association between weekend catch-up sleep and lower probability of cognitive decline.^10^ However, excessive weekend catch-up sleep may disrupt circadian rhythms in sleep-wake patterns (known as social jetlag) and thus not confer additional benefits for brain health. As higher day-to-day variability in sleep schedules has previously been associated with vascular risk factors^11,12^ and increased dementia risk,^13^ the relation of weekend catch-up sleep with dementia remains controversial. Importantly, most existing studies employed cross-sectional designs or relied on self-reported sleep measures prone to reporting bias, which limit the ability to draw causal inferences about the potential protective effects of compensating for sleep debt versus the harms associated with circadian disruption.

To address the roles of these two potentially competing mechanisms of weekend catch-up sleep, the current study utilizes objectively measured sleep durations to investigate the association between weekend catch-up and incident dementia. We hypothesized that moderate weekend catch-up sleep was associated with lower dementia risk, particularly among individuals with less sleep on weekdays.

## Methods

### Study population

This study was based on the UK Biobank, a population-based cohort study commenced in 2006-2010. The UK Biobank recruited over 500,000 residents across the United Kingdom aged 37 years and older at 22 assessment centres and collected extensive genetic and phenotypic data.^14,15^ Between 2013-2015, a random subset was invited to participate in a 3-axis logging accelerometer sub-study for seven days.^16^ The protocol of UK Biobank was approved by the North West Multi-Centre Research Ethics Committee (REC reference: 11/NW/0382), and all participants provided informed consent before data collection.

Among the 103,684 participants who wore the accelerometer, we included participants with ≥3 valid wearing days (>16 hours/day and at least one weekend day),^17^ aged 50 years or older at the time of accelerometery,^18^ and free of all-cause dementia before and in two years after participating in the sub-study to reduce the risk of reverse causality.^19^ We further excluded participants who had extreme weekday sleep duration, weekend sleep duration, or weekday-weekend difference (<0.5^th^ or >99.5^th^ percentiles). The final sample size was 83,776 (**Figure 1**).

**Figure 1.**
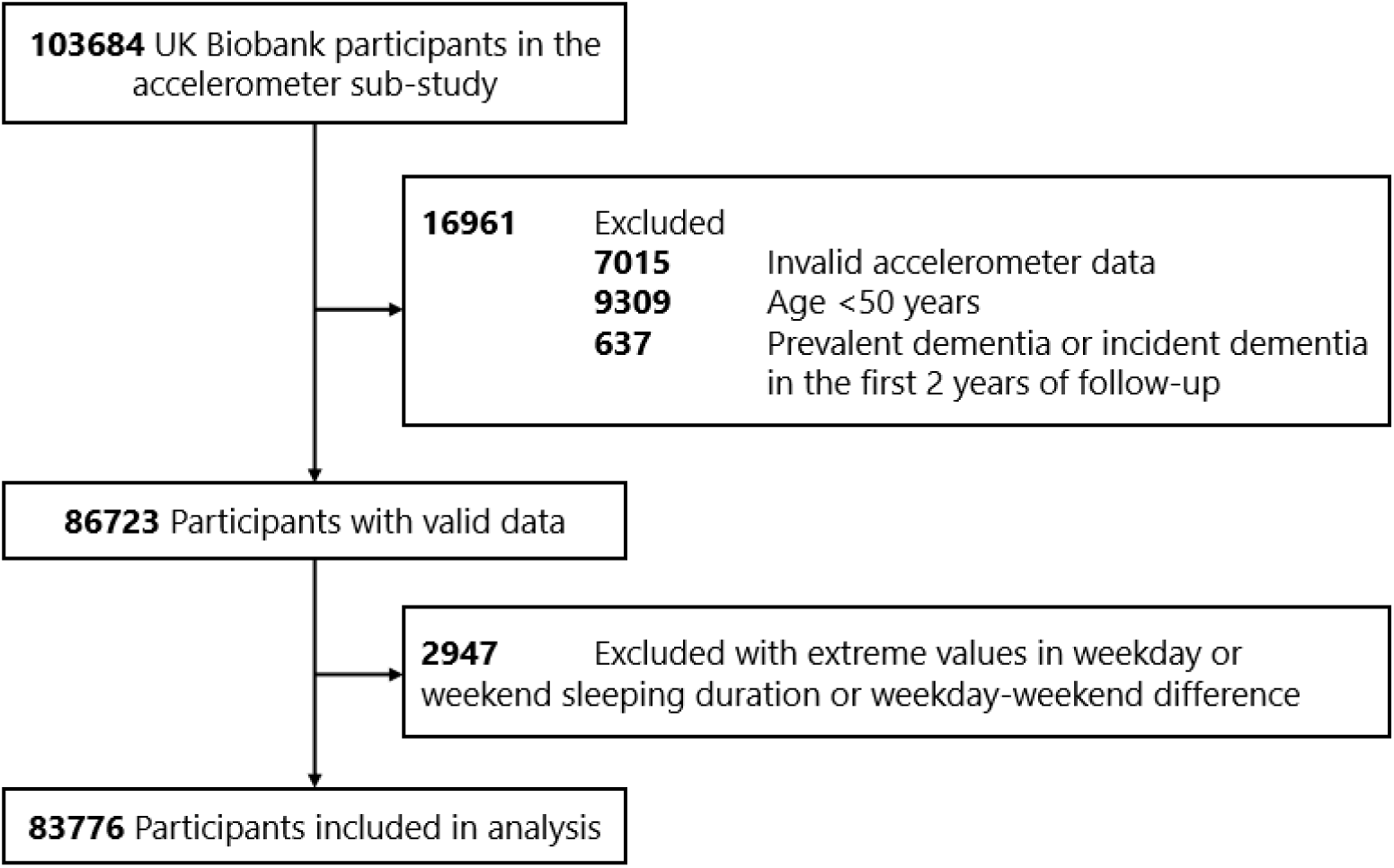
Participants inclusion flow chart

### Accelerometer-derived sleep duration

The participants were invited to wear the Axivity AX3 triaxial accelerometer on their dominant wrist for seven days,^20^ and mail the device back to the coordinating centre after the seven-day period.^16^ Daily sleep duration was extracted from raw accelerometer data using a previously published machine-learning algorithm for the UK Biobank.^21^ In a validation study (n=152), the algorithm showed high classification performance (accuracy 88%) in identifying movement behaviours, and the precision for identifying sleeping behaviour was >95%.^21^

We calculated the average sleep duration for weekday (Monday to Friday) and weekend (Saturday and Sunday), respectively, and calculated the weekend-weekday difference as weekend catch-up sleep.^22^ According to the study population distribution, we categorized the weekend catch-up sleep duration as both multi-class (≤0.5, >0.5-1, >1-1.5, >1.5-2, or >2 hours, as the primary exposure) and binary variables (≤0.5 or >0.5 hour, as the secondary exposure). We specified 0.5 hour as the cut-off because it was close to the population median (0.41 hour) and conservatively identifies the catch-up population.^22^ We also included the weekend-weekday difference as a continuous variable to assess the linear and non-linear associations.

### Dementia ascertainment

The outcome of interest in this study was incident all-cause dementia. We used a linkage-based algorithm to define incident dementia, combining information from electronic health records (EHRs) in primary care, hospital admissions and death registry, as was described previously.^23^ We used the date of the first occurrence in any of the abovementioned sources as the diagnosis date of dementia, which showed a positive predictive value of 82.5%.^23^

### Covariates

We included multiple covariates for confounding adjustments based on the existing literature.^17^ Demographic factors and socioeconomic status included age at accelerometer assessment, sex, race and ethnicity (white or non-white), and Townsend deprivation index (as tertiles, reflecting social deprivation level). Lifestyle factors included BMI category (under or normal weight [<25.0 kg/m²] /overweight [25.0 −29.9 kg/m²] /obesity [≥30.0 kg/m²]), smoking status (never/former/current), alcohol drinking status (never/former/current), physical activity (measured by the International Physical Activity Questionnaire, in metabolic equivalent hours per week), sleep duration on weekday (hours), shift work status, and chronotypes defined as a previous study.^24^ Health conditions included diabetes, high blood pressure, depression, angina, stroke, and heart attack updated until accelerometer wearing, and depression status defined according to 2-item Patient Health Questionnaire collected at recruitment,^25^ which are potential confounders.

### Statistical analysis

We described baseline characteristics of the participants using means (standard deviations, SDs) for continuous variables and numbers (percentages) for categorical variables. Missing values of covariates were imputed using multiple imputation with chained equations.^26^

We used Cox proportional hazard regression models to assess the association between accelerometer-measured weekend catch-up sleep and incident dementia. Weekend catch-up sleep was included as multi-class, continuous, and binary variables, in separate models. Person-time was calculated from the accelerometer-wearing date to the diagnosis of dementia, the ascertainment of death, or the end of follow-up (December 2022), whichever came first. The hazard ratios (HRs) and confidence intervals (CIs) of incident dementia were sequentially adjusted for age at accelerometer assessment and sex in Model 1, ethnicity, Townsend deprivation index, BMI category, smoking status, alcohol drinking status, physical activity, sleep duration on weekday, shift work status, and chronotype in Model 2, and diabetes, high blood pressure, depression, angina, stroke, heart attack, and depression in Model 3. To evaluate potential non-linear patterns in the association, we modelled weekend catch-up sleep as a continuous variable using restricted cubic splines (RCSs), setting the median as the reference point and determining the optimal degrees of freedom based on the maximum Akaike information criterion.^27^

To further assess whether the association differed by weekday sleep duration, we stratified the analysis by weekday sleep duration (<8 or ≥8 hours) according to the population distribution (only 4,637 [5.5%] participants had weekday sleep duration <7 hours, which led to model convergence failure). We also preplanned other stratified analyses to assess the associations in subgroups of participants defined by age (<65 years or ≥65 years), sex, Townsend deprivation index (below or above median), BMI (obesity or non-obesity), and smoking status (never or ever). We used likelihood ratio tests to evaluate the statistical significance of the corresponding multiplicative interaction term in the Cox regression models, comparing models with and without the interaction term.

In a secondary analysis, we explored the joint association of weekday sleep duration and weekend catch-up sleep by categorizing participants into four groups based on a 2×2 classification. We further classified participants into three categories: (1) average sleep <8 hours, (2) average sleep ≥8 hours without weekend catch-up sleep (≤0.5 hour), and (3) average sleep ≥8 hours with weekend catch-up sleep (>0.5 hour). To examine the potential differential associations of weekday and weekend sleep durations with dementia, we assessed the relationships of average sleep duration, weekday sleep duration, and weekend sleep duration with incident all-cause dementia, using the same approaches as specified above. When evaluating the independent associations of weekday and weekend sleep durations, we mutually adjusted for each in the same model.

We conducted several sensitivity analyses to verify the robustness of the primary findings. We first excluded participants with baseline cardiovascular diseases or diabetes, who might have altered sleeping behaviour because of the disease status. Secondly, we excluded participants with depression at baseline because sleeping disorder is often a manifestation of depression. Thirdly, we excluded participants who developed dementia within the first 5 years of follow-up to further reduce the impact of reverse causality. Finally, we specified participants with weekend-weekday difference ≤0 hours as the reference group and reassessed the association.

All analyses were performed using R version 4.3.0. Statistical significance level was set to be two-sided *P*-values <0.05. We do not correct the *P*-values for multiplicity in secondary and subgroup analyses given their exploratory nature.

## Results

### Baseline characteristics

Among the 83,776 participants, the mean age at the time of accelerometer assessment was 63.3 years (SD: 6.7), 56.5% were female, and the mean weekend catch-up sleep duration was 0.49 hours (SD: 1.21, **Table 1**). Among the study participants, 44,520 had none or ≤0.5-hour weekend catch-up group (53.1%), of whom 29,413 (35.1%) had a weekend-weekday difference in sleep duration ≤0 hour. Participants engaging in longer weekend catch-up sleep were younger, less likely to be female, and had shorter sleep duration on weekdays (mean: 8.4 hours, SD: 1.1 for >0.5-hour catch-up group), compared to those with none or ≤0.5 hour of catch-up sleep (mean: 9.0 hours, SD: 1.2). **Table S1** showed the mean sleep durations on each day of week.

**Table 1.** Baseline characteristics of participants.

| Variable | Overall | Weekend catch-up sleep |  |  |  |  |
| --- | --- | --- | --- | --- | --- | --- |
|  |  | None or ≤0.5 hour | >0.5-1 hour | >1-1.5 hours | >1.5-2 hours | >2 hours |
| N | 83776 | 44520 | 13969 | 10150 | 6458 | 8679 |
| Age at accelerometer assessment (mean (SD)) | 63.3 (6.7) | 64.5 (6.4) | 63.4 (6.6) | 62.4 (6.7) | 61.3 (6.7) | 59.9 (6.4) |
| Male (%) | 36478 (43.5) | 18822 (42.3) | 6159 (44.1) | 4444 (43.8) | 2962 (45.9) | 4091 (47.1) |
| Townsend deprivation index (mean (SD)) | 14.5 (11.7) | 14.3 (11.6) | 14.1 (11.3) | 14.5 (11.7) | 15.1 (12.2) | 15.8 (12.4) |
| College or university education (%) | 35667 (42.6) | 18528 (41.6) | 6200 (44.4) | 4542 (44.7) | 2766 (42.8) | 3631 (41.8) |
| White ethnicity (%) | 81565 (97.4) | 43477 (97.7) | 13629 (97.6) | 9853 (97.1) | 6262 (97.0) | 8344 (96.1) |
| BMI category (%) |  |  |  |  |  |  |
| -Underweight | 2264 (2.7) | 1209 (2.7) | 393 (2.8) | 271 (2.7) | 163 (2.5) | 228 (2.6) |
| -Normal weight | 30313 (36.2) | 16210 (36.4) | 5222 (37.4) | 3692 (36.4) | 2310 (35.8) | 2879 (33.2) |
| -Overweight | 35016 (41.8) | 18566 (41.7) | 5846 (41.8) | 4298 (42.3) | 2709 (41.9) | 3597 (41.4) |
| -Obesity | 16183 (19.3) | 8535 (19.2) | 2508 (18.0) | 1889 (18.6) | 1276 (19.8) | 1975 (22.8) |
| Smoking status (%) |  |  |  |  |  |  |
| -Never | 47382 (56.6) | 24952 (56.0) | 7965 (57.0) | 5806 (57.2) | 3687 (57.1) | 4972 (57.3) |
| -Former | 30966 (37.0) | 16792 (37.7) | 5136 (36.8) | 3674 (36.2) | 2352 (36.4) | 3012 (34.7) |
| -Current | 5428 (6.5) | 2776 (6.2) | 868 (6.2) | 670 (6.6) | 419 (6.5) | 695 (8.0) |
| Alcohol drinking status (%) |  |  |  |  |  |  |
| -Never | 2424 (2.9) | 1310 (2.9) | 391 (2.8) | 289 (2.8) | 159 (2.5) | 275 (3.2) |
| -Former | 2251 (2.7) | 1195 (2.7) | 351 (2.5) | 280 (2.8) | 172 (2.7) | 253 (2.9) |
| -Current | 79101 (94.4) | 42015 (94.4) | 13227 (94.7) | 9581 (94.4) | 6127 (94.9) | 8151 (93.9) |
| Physical activity – MET-Minutes/week (mean (SD)) | 2538.5 (2438.5) | 2583.4 (2448.9) | 2524.2 (2431.7) | 2490.5 (2408.4) | 2445.7 (2399.1) | 2457.0 (2455.3) |
| Sleep duration on weekday – Hours/day (mean (SD)) | 8.7 (1.2) | 9.0 (1.2) | 8.6 (1.1) | 8.4 (1.1) | 8.4 (1.2) | 8.3 (1.2) |
| Sleep duration on weekend – Hours/day (mean (SD)) | 9.2 (1.4) | 8.6 (1.2) | 9.3 (1.1) | 9.7 (1.1) | 10.1 (1.2) | 11.1 (1.4) |
| Diabetes (%) | 2372 (2.8) | 1297 (2.9) | 341 (2.4) | 279 (2.7) | 194 (3.0) | 261 (3.0) |
| High blood pressure (%) | 61461 (73.4) | 33045 (74.2) | 10231 (73.2) | 7389 (72.8) | 4609 (71.4) | 6187 (71.3) |
| | | None or $\leq 0.5$ hour | >0.5-1 hour | >1-1.5 hours | >1.5-2 hours | >2 hours |
| N | 83776 | 44520 | 13969 | 10150 | 6458 | 8679 |
| Depression (%) | 3043 (3.6) | 1601 (3.6) | 464 (3.3) | 335 (3.3) | 243 (3.8) | 400 (4.6) |
| Angina (%) | 1896 (2.3) | 1052 (2.4) | 338 (2.4) | 195 (1.9) | 139 (2.2) | 172 (2.0) |
| Stroke (%) | 854 (1.0) | 483 (1.1) | 153 (1.1) | 85 (0.8) | 60 (0.9) | 73 (0.8) |
| Heart attack (%) | 1354 (1.6) | 740 (1.7) | 227 (1.6) | 130 (1.3) | 114 (1.8) | 143 (1.6) |

SD: standard deviation, BMI: Body mass index.

### Accelerometer-measured weekend catch-up sleep and incident dementia

During a total of 667,928 person-years (median follow-up = 8.0 years), 713 participants developed all-cause dementia. We observed a significant lower risk of dementia among individuals with a moderate level of weekend catch-up sleep (1-1.5 hours/day). The fully adjusted HRs and 95%CIs across increasing categories of weekend catch-up sleep (none or ≤0.5, >0.5-1, >1-1.5, >1.5-2, and >2 hours) were 1 (reference), 0.91 (0.74-1.11), 0.64 (0.49-0.86), 0.84 (0.60-1.16), and 0.83 (0.60-1.16), respectively (**Table 2**). **Figure S1** illustrated the association between weekend catch-up sleep duration and the risk of incident dementia, modelled using a RCS. The analysis demonstrated a generally decreasing hazard for dementia with increasing weekend catch-up sleep duration, with the lowest risk around 1-1.5 hours of additional weekend sleep, despite of lacking statistical non-linearity (*P*-value=0.563).

**Table 2.** Hazard ratios of incident dementia according to accelerometer-measured weekend catch-up sleep.

| **Weekend catch-up sleep** | **N** | **Dementia Incidence** | **Model 1** | **Model 2** | **Model 3** |
| --- | --- | --- | --- | --- | --- |
| None or $\leq 0.5$ hour | 44520 | 462/354120 | 1.00 (Reference) | 1.00 (Reference) | 1.00 (Reference) |
| >0.5-1 hour | 13969 | 118/111199 | 0.93 (0.76-1.14) | 0.91 (0.74-1.11) | 0.91 (0.74-1.11) |
| >1-1.5 hours | 10150 | 54/81260 | **0.67 (0.50-0.88)** | **0.64 (0.49-0.85)** | **0.64 (0.49-0.86)** |
| >1.5-2 hours | 6458 | 39/51750 | 0.88 (0.64-1.22) | 0.85 (0.61-1.18) | 0.84 (0.60-1.16) |
| >2 hours | 8679 | 40/69600 | 0.89 (0.65-1.24) | 0.84 (0.61-1.16) | 0.83 (0.60-1.16) |

Model 1 was adjusted for age at accelerometer assessment and sex. Model 2 was based on model 1 and further adjusted for ethnicity, Townsend deprivation index, BMI category, smoking status, alcohol drinking status, physical activity, sleep duration on weekday, shift work status, and chronotype. Model 3 was based on model 2 and further adjusted for diabetes, high blood pressure, depression, angina, stroke, heart attack, and depression.

When we further stratified the analyses by average weekday sleep duration, the associations were stronger among participants with less weekday sleep duration (**Table 3** and **Figure 2**). In the RCS analysis, the likelihood ratio test yielded a *P*-interaction of 0.039, indicating a statistically significant interaction. For participants with a weekday sleep duration <8 hours (**Figure 3A**), the association was approximately linear (*P*-linear=0.003, *P*-nonlinear=0.449), with the fully adjusted HRs and 95%CIs across increasing categories of weekend catch-up sleep (none or ≤0.5, >0.5-1, >1-1.5, and >1.5 hours) being 1 (reference), 0.97 (0.69-1.37), 0.49 (0.29-0.81), and 0.52 (0.34-0.82), respectively. Participants with >0.5 hour of catch-up sleep had 0.69-fold (0.52-0.91, *P*-value=0.009) risk of dementia compared with those with none or ≤0.5 hour, and the HR of per hour increase in weekend catch-up sleep was 0.75 (95% CI: 0.65-0.87, *P*-value<0.001, **Table 3**). On the contrary, there was no significant association for participants with a weekday sleep duration ≥8 hours (*P*-linear=0.362, *P*-nonlinear=0.265), though associations remained in the same direction for >1-1.5 hours (**Figure 3B**).

**Figure 2.**
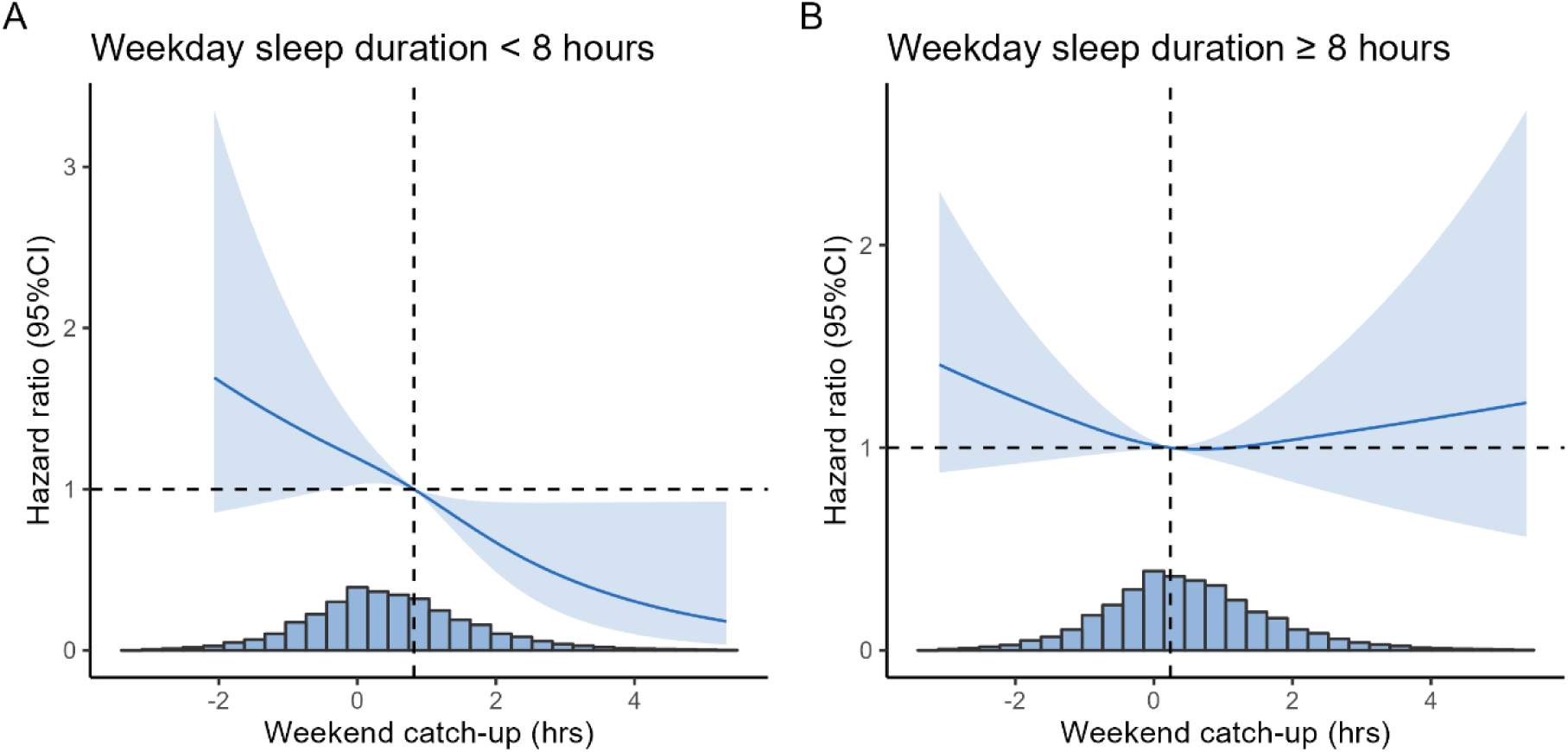
*Associations of accelerometer-measured weekend catch-up sleep with risk of incident dementia modelled using restricted cubic spline model by weekday sleep duration* Weekend catch-up sleep was defined as the weekend-weekday sleep duration difference. The Cox model was adjusted for age at accelerometer assessment, sex, education, ethnicity, Townsend deprivation index, BMI category, smoking status, alcohol drinking status, physical activity, diabetes, high blood pressure, depression, angina, stroke, heart attack, and depression. Reference point was set to be population median sleep duration on x-axis. Number of knots was chosen based on AIC criteria. For participants with weekday sleep duration <8 hours (A), *P*-linear=0.003, *P*-nonlinear=0.449. For participants with weekday sleep duration ≥8 hours (B), *P*-linear=0.362, *P*-nonlinear=0.265. The likelihood ratio test for the interaction term yielded a Chi-square statistic of 6.46 (degrees of freedom = 2), with a *P*-value of 0.039, indicating a statistically significant interaction.

**Table 3.** Hazard ratios of incident dementia by categories accelerometer-measured weekend catch-up sleep by weekday sleep duration.

| Variable | Weekday sleep duration <8 hrs | P-value | Weekday sleep duration ≥8 hrs | P-value | P for difference |
| --- | --- | --- | --- | --- | --- |
| **By duration** |  |  |  |  |  |
| None or ≤0.5 hour | 1.00 (Reference) |  | 1.00 (Reference) |  |  |
| >0.5-1 hour | 0.97 (0.69-1.37) | 0.871 | 0.85 (0.66-1.10) | 0.230 | 0.545 |
| >1-1.5 hours | **0.49 (0.29-0.81)** | **0.006** | 0.72 (0.51-1.01) | 0.059 | 0.221 |
| >1.5 hours | **0.52 (0.34-0.82)** | **0.004** | 1.01 (0.76-1.35) | 0.921 | **0.013** |
| **Per hour** | **0.75 (0.65-0.87)** | **<0.001** | 0.96 (0.89-1.04) | 0.356 | **0.003** |
| **Binary** |  |  |  |  |  |
| None or ≤0.5 hour | 1.00 (Reference) |  | 1.00 (Reference) |  |  |
| >0.5 hour | **0.69 (0.52-0.91)** | **0.009** | 0.86 (0.72-1.04) | 0.125 | 0.197 |

The Cox model was adjusted for age at accelerometer assessment, sex, education, ethnicity, Townsend deprivation index, BMI category, smoking status, alcohol drinking status, physical activity, diabetes, high blood pressure, depression, angina, stroke, heart attack, and depression. *P* for difference was calculated by comparing the difference of hazards ratios between weekday sleep duration <8 hours and ≥8 hours.

### Secondary analysis

When we dichotomized the participants according to the weekend catch-up sleep duration, participants with >0.5 hour of catch-up sleep had 0.81-fold (0.70-0.95, *P*-value=0.010) risk of dementia compared with those with none or ≤0.5 hour. When measured as a continuous variable, each hour increment in weekend catch-up sleep was associated with a 9% lower risk of dementia (HR: 0.91, 95% CI: 0.84-0.98, *P*-value=0.008, **Table S2**).

We further categorized participants by overall sleep duration and presence of weekend catch-up sleep (**Table S3**). Compared to short sleepers whose overall sleep duration <8 hours, those who had overall sleep duration ≥8 hours, both with (HR = 0.67, 95% CI: 0.55–0.82, *P*-value<0.001) and without (HR = 0.77, 95% CI: 0.64–0.92, *P*-value=0.004) weekend catch-up, were at lower risk of dementia.

These findings were confirmed by the joint associations of weekday sleep and weekend catch-up (**Table S3**). Compared to participants with weekday sleep <8 hours and without weekend catch-up, those with weekday sleep <8 hours who had weekend catch-up showed an HR of 0.74 (95% CI: 0.56–0.98, *P*-value=0.032). Compared to those who slept <8 hours without weekend catch-up, participants who slept ≥8 hours on weekday, either with (HR = 0.62, 95% CI: 0.49-0.79, *P*-value<0.001) or without (HR = 0.72, 95% CI: 0.59-0.89, *P*-value=0.003) weekend catch-up, had lower dementia risk.

We also explored the potential associations of sleep durations on different days with risk of dementia, and the associations for average, weekday, and weekend sleep durations all showed non-linear patterns (**Table S4** and **Figure S2)**. Compared with participants with a daily sleep duration of >8-9 hours, those who had less sleep were at significantly higher dementia risk, with HR being 2.05 (95% CI: 1.49-2.82) for ≤7 hours and 1.25 (95% CI: 1.02-1.54) for >7-8 hours. This U-shape trend was both observed for weekday and weekend sleep durations.

### Subgroup and sensitivity analysis

The associations between weekend catch-up sleep and incident dementia were similar across the subgroups defined by the sociodemographic and lifestyle factors (**Table S5** and **Figure S3**). Although the association for >1-1.5 hours catch-up sleep tended to be stronger in adults aged ≥65 years (HR = 0.38, 95% CI: 0.15-0.96) than in those aged 50-64 years (HR = 0.66, 95% CI: 0.49-0.88), the interaction was not statistically significant (*P*-interaction = 0.568).

The associations were also robust in several sensitivity analyses (**Table S6**). The HR (95%CI) for >1-1.5 hours of catch-up sleep was 0.64 (95% CI: 0.47-0.88) when we excluded participants with cardiovascular diseases or diabetes, 0.60 (95% CI: 0.44-0.80) when we excluded participants with depression, and 0.64 (95% CI: 0.48-0.85) when we excluded dementia cases in the first 5-year follow-up. When using catch-up of ≤0 hour as the reference group, the associations were similar (**Table S7**).

## Discussion

In this prospective cohort study, we found that moderate weekend catch-up sleep was associated with a lower risk of incident all-cause dementia in later life. Participants with 1-1.5 hours of weekend catch-up sleep had the most pronounced reduction in dementia risk. The association was stronger among individuals with weekday sleep <8 hrs/d. Similar associations were observed across the subgroups defined by the sociodemographic and lifestyle factors.

### Comparison with other studies and possible explanations

Our findings add to the growing evidence on sleep pattern and cognitive health. Prior studies have consistently demonstrated that inadequate sleep is associated with higher dementia risk, highlighting the importance of healthy sleep in brain health. A previous meta-analysis reported a U-curve association for sleep duration and dementia with a turning point at 7-8 hours,^5^ which was further confirmed by another study in the UK Biobank^28^ and this study. However, previous studies primarily based on self-reported sleep duration and focused on average sleep duration, with limited attention to the weekday-weekend variations in sleep. Using data from objective, unbiased sleep measurements, the current study offers new evidence on the potentially beneficial association between compensatory sleep and dementia. Our findings echoed a cross-sectional study among older adults in Taipei city, which used sleep diaries and accelerometer to assess sleep duration and reported a 73–74% lower odds of cognitive dysfunction among individuals with weekend catch-up sleep.^10^ The present results indicate that not only maintaining adequate sleep on weekdays, but weekend compensatory catch-up sleep may also play a protective role. If proven causal, these findings could contribute to the existing recommendations for dementia prevention.^29^ Our findings also indicated that the association was stronger among participants with weekday sleep duration <8 hours, which suggests that moderate catch-up sleep may primarily act as a compensatory mechanism. Those with weekday sleep ≥8 hours likely already experience optimal cognitive and physiological benefits, leaving little room for further compensation.

Additionally, our results suggest that the lowest risk was observed among individuals who achieved moderate levels of weekend catch-up sleep (1-1.5 hours), although the non-linearity did not reach statistical significance. This may stem from biological and lifestyle factors associated with both sleep and brain health. Existing studies have explored how adequate sleep supports long-term brain functions, such as neural repair, synaptic plasticity, and waste clearance through the glymphatic system,^30–32^ which are essential for preventing neurodegenerative processes associated with dementia. Moderate catch-up sleep might restore certain cognitive and physiological processes affected by sleep debt,^33^ such as improved glymphatic clearance^34^ of neurotoxic waste,^30^ which has been shown to occur during deep sleep. However, longer durations of weekend sleep could signal a more chronic sleep imbalance, disrupted circadian rhythms, or other health conditions that independently contribute to dementia risk. Although there is no universal optimal range, excessive weekend catch-up may also indicate fragmented sleep or lifestyle stressors during the week that cannot be fully mitigated by longer weekend rest, suggesting diminishing returns beyond a certain threshold. Further research is needed to elucidate the underlying mechanisms and to identify the optimal range of weekend catch-up sleep, potentially varying by weekday sleep duration, that supports cognitive resilience and informs targeted interventions.

### Strengths and limitations

This study has several strengths, including its large sample size, prospective design, and the objective measurements for sleep duration with a higher precision compared with self-reported sleep duration. However, some limitations should be noted. First, the accelerometer data may well capture sleep duration, but we did not include information on sleep quality or identify various sleep characteristics such as wake after sleep onset, sleep efficiency, sleep timing and regularity. Although our findings are comparable with estimates from previous studies,^21^ the sleep measurements in this study may include time spent in bed, potentially leading to an overestimation of overall sleep duration, which needs further calibration using polysomnographic measurements. Residual or unmeasured confounding could still bias the findings. In addition, while accelerometer data provided detailed seven-day sleep patterns, it was collected at a single time point, and we were unable to further investigate the associations of long-term and time-varying sleep behaviors. Considering the long-term preclinical phase of dementia, this warrants further investigations.^1^ Furthermore, the cohort primarily included adults of White ethnicity, which may limit the generalizability of findings to more diverse populations. Finally, the respondents to this sub-study of the UK Biobank could be healthier than the general population, which may introduce selection bias as they could be less susceptible to dementia, although this may not fully diminish its representativeness in assessing exposure-disease relationships.^35^

## Conclusions

In summary, weekend catch-up sleep was associated with lower risk of dementia among middle-aged and older adults. Our findings suggest that moderate weekend catch-up sleep, particularly around 1–1.5 hours, may be associated with the lowest dementia risk. The association was stronger among individuals with less sleep on weekdays. Future studies are needed to confirm the findings and determine the optimal sleep compensation strategies for dementia prevention.

## Data Availability

Data can be shared through mechanisms detailed at https://www.ukbiobank.ac.uk/.

## Author information

### Contributors

HC, TH and CY made a substantial contribution to the concept of the the study design. HC and TS drafted the manuscript and conducted statistical analysis. TH and CY provided administrative, technical, or material support and offered supervision. All authors contributed to interpretation of data and had provided critical revision of the manuscript for important intellectual content. TH and CY have access to all the data used in this study and take full responsibility to this study. The corresponding author attests that all listed authors meet authorship criteria and that no others meeting the criteria have been omitted. CY is the guarantor.

## Ethical approval

The North West Multi-Centre Research Ethics Committee approved the protocol of UK Biobank (REC reference: 11/NW/0382), and all participants provided informed consent before data collection.

## Data sharing

Data can be shared through mechanisms detailed at https://www.ukbiobank.ac.uk/.

## Transparency

The manuscript’s guarantor affirms that the manuscript is an honest, accurate, and transparent account of the study being reported; that no important aspects of the study have been omitted; and that any discrepancies from the study as planned (and, if relevant, registered) have been explained.

## Dissemination to participants and related patient and public communities

The results of the research will be disseminated to the public through broadcasts, newspaper, television, social media, conference, and popular science articles.

