## Supplemental tables and figures for "Accelerometer-measured weekend catch-up sleep and incident dementia: a prospective cohort study"

### Supplement Material

*Table S 1 Mean sleep durations at each day of week*

| Day | Mean (SD) |
|---|---|
| Monday | 8.83 (1.62) |
| Tuesday | 8.63 (1.56) |
| Wednesday | 8.69 (1.61) |
| Thursday | 8.71 (1.64) |
| Friday | 8.72 (1.67) |
| Saturday | 9.08 (1.65) |
| Sunday | 9.33 (1.65) |
| Average | 8.86 (1.14) |
| Weekday | 8.71 (1.20) |
| Weekend | 9.21 (1.43) |
| Weekend - weekday | 0.49 (1.21) |

*Table S 2 Hazard ratios of incident dementia by categories accelerometer-measured weekend catch-up sleep (per hour and binary variable)*

| Weekend catch-up sleep | N | Dementia Incidence | Model 1 | Model 2 | Model 3 |
|---|---|---|---|---|---|
| **Per hour** | 83776 | 713/667928 | 0.93 (0.87-1.00) | **0.91 (0.85-0.98)** | **0.91 (0.84-0.98)** |
| **Binary** | | | | | |
| None or ≤0.5 hour | 44520 | 462/354120 | 1.00 (Reference) | 1.00 (Reference) | 1.00 (Reference) |
| >0.5 hour | 39256 | 251/313809 | **0.84 (0.72-0.99)** | **0.82 (0.70-0.95)** | **0.81 (0.70-0.95)** |

Model 1 was adjusted for age at accelerometer assessment and sex. Model 2 was based on model 1 and further adjusted for ethnicity, Townsend deprivation index, BMI category, smoking status, alcohol drinking status, physical activity, sleep duration on weekday, shift work status, and chronotype. Model 3 was based on model 2 and further adjusted for diabetes, high blood pressure, depression, angina, stroke, heart attack, and depression.

*Table S 3 Hazard ratios of incident dementia by joint categories accelerometer-measured average sleep durations and weekend catch-up sleep*

| Category | N | Case/PY | Hazard Ratio (95%CI) | *P*-value |
|---|---|---|---|---|
| **Joint category of overall sleep duration and weekend catch-up** | | | | |
| Overall sleep duration <8 hrs | 19099 | 195/152347.0 | 1.00 (Reference) | |
| Overall sleep duration ≥8 hrs - No catch-up | 34437 | 329/273908.3 | **0.77 (0.64-0.92)** | **0.004** |
| Overall sleep duration ≥8 hrs - Catch-up | 30240 | 189/241672.8 | **0.67 (0.55-0.82)** | **<0.001** |
| **Joint category of weekday sleep duration and weekend catch-up** | | | | |
| Weekday sleep duration <8 hrs - No catch-up | 8952 | 116/71227.3 | 1.00 (Reference) | |
| Weekday sleep duration <8 hrs- Catch-up | 14517 | 89/116273.2 | **0.74 (0.56-0.98)** | **0.032** |
| Weekday sleep duration ≥8 hrs - No catch-up | 35568 | 346/282892.2 | **0.72 (0.59-0.89)** | **0.003** |
| Weekday sleep duration ≥8 hrs - Catch-up | 24739 | 162/197535.3 | **0.62 (0.49-0.79)** | **<0.001** |

The Cox model was adjusted for age at accelerometer assessment, sex, education, ethnicity, Townsend deprivation index, BMI category, smoking status, alcohol drinking status, physical activity, diabetes, high blood pressure, depression, angina, stroke, heart attack, and depression.

*Table S 4 Hazard ratios of incident dementia by accelerometer-measured average sleep duration and sleep durations on weekday and weekend*

| Duration (hrs) | Average sleep | Weekday sleep | Weekend sleep |
|---|---|---|---|
| 0-7 | **2.05 (1.49-2.82)** | **1.42 (1.02-1.98)** | **1.41 (1.02-1.95)** |
| >7-8 | **1.25 (1.02-1.54)** | 1.03 (0.84-1.28) | 1.18 (0.94-1.47) |
| >8-9 | 1.00 (Reference) | 1.00 (Reference) | 1.00 (Reference) |
| >9 | 1.01 (0.85-1.20) | 1.03 (0.85-1.24) | 0.90 (0.75-1.09) |

The Cox model was adjusted for age at accelerometer assessment, sex, education, ethnicity, Townsend deprivation index, BMI category, smoking status, alcohol drinking status, physical activity, diabetes, high blood pressure, depression, angina, stroke, heart attack, and depression. The HRs for weekday and weekend sleep was mutually adjusted in addition to these covariates.

*Table S 5 Hazard ratios of incident dementia by accelerometer-measured weekend catch-up sleep (multiclass) in study subgroups*

| Subgroup | Case/PY | None or ≤0.5 hour | >0.5-1 hour | >1-1.5 hours | >1.5-2 hours | >2 hours | *P*-interaction |
|---|---|---|---|---|---|---|---|
| Age | | | | | | | 0.568 |
| <65 | 83/368875.4 | 1.00 (Reference) | 0.85 (0.68, 1.05) | **0.66 (0.49, 0.88)** | 0.80 (0.56, 1.15) | 0.72 (0.49, 1.05) | |
| ≥65 | 630/299052.7 | 1.00 (Reference) | 1.14 (0.65, 2.00) | **0.38 (0.15, 0.96)** | 0.72 (0.32, 1.63) | 0.73 (0.37, 1.45) | |
| Sex | | | | | | | 0.235 |
| Women | 334/378498.2 | 1.00 (Reference) | 0.85 (0.62, 1.16) | 0.79 (0.53, 1.16) | 1.17 (0.75, 1.82) | 0.88 (0.54, 1.45) | |
| Men | 379/289429.9 | 1.00 (Reference) | 0.95 (0.73, 1.24) | **0.53 (0.35, 0.79)** | 0.61 (0.37, 1.01) | 0.80 (0.52, 1.23) | |
| Social deprivation | | | | | | | 0.401 |
| High | 352/333070.4 | 1.00 (Reference) | 0.89 (0.67, 1.20) | 0.68 (0.46, 1.01) | 0.87 (0.56, 1.37) | 0.79 (0.50, 1.25) | |
| Low | 361/334857.7 | 1.00 (Reference) | 0.92 (0.69, 1.21) | **0.60 (0.40, 0.91)** | 0.81 (0.50, 1.32) | 0.88 (0.55, 1.41) | |
| BMI | | | | | | | 0.507 |
| Normal/Underweight | 266/260654.6 | 1.00 (Reference) | 1.04 (0.75, 1.44) | 0.74 (0.47, 1.17) | 1.14 (0.68, 1.91) | 1.27 (0.74, 2.17) | |
| Overweight/Obesity | 447/407273.5 | 1.00 (Reference) | 0.83 (0.64, 1.08) | **0.59 (0.41, 0.85)** | 0.70 (0.46, 1.08) | 0.69 (0.45, 1.04) | |
| Smoking | | | | | | | 0.246 |
| Ever smoking | 378/288965.0 | 1.00 (Reference) | 0.96 (0.73, 1.26) | **0.62 (0.42, 0.92)** | 0.61 (0.37, 1.02) | 0.69 (0.43, 1.12) | |
| Never smoking | 335/378963.1 | 1.00 (Reference) | 0.85 (0.62, 1.15) | 0.67 (0.44, 1.00) | 1.12 (0.73, 1.73) | 1.00 (0.64, 1.57) | |

The Cox model was adjusted for age at accelerometer assessment, sex, education, ethnicity, Townsend deprivation index, BMI category, smoking status, alcohol drinking status, physical activity, diabetes, high blood pressure, depression, angina, stroke, heart attack, and depression. The stratification variables were not adjusted for the corresponding analyses. *P*-interactions were calculated from the likelihood ratio test comparing the model with and without the interaction term crossing weekend catch-up sleep and the stratification variable.

*Table S 6 Hazard ratios of incident dementia by accelerometer-measured weekend catch-up sleep in sensitivity analyses*

| Variable | Excluding participants with cardiovascular diseases or diabetes | Excluding participants with depression | Excluding cases in the first 5 years |
|---|---|---|---|
| **By duration** | | | |
| None or ≤0.5 hour | 1.00 (Reference) | 1.00 (Reference) | 1.00 (Reference) |
| >0.5-1 hour | 0.94 (0.75-1.17) | 0.91 (0.74-1.11) | 0.91 (0.74-1.11) |
| >1-1.5 hours | **0.64 (0.47-0.88)** | **0.60 (0.44-0.80)** | **0.64 (0.48-0.85)** |
| >1.5-2 hours | 0.85 (0.59-1.23) | 0.81 (0.57-1.13) | 0.84 (0.60-1.16) |
| >2 hours | 0.92 (0.64-1.31) | **0.89 (0.83-0.96)** | **0.91 (0.84-0.98)** |
| **Per hour** | 0.93 (0.86-1.01) | **0.80 (0.68-0.94)** | **0.81 (0.70-0.95)** |
| **Binary** | | | |
| None or ≤0.5 hour | 1.00 (Reference) | 1.00 (Reference) | 1.00 (Reference) |
| >0.5 hour | 0.84 (0.71-1.00) | **0.80 (0.68-0.94)** | **0.81 (0.70-0.95)** |

The sensitivity analyses were based on Cox model adjusted for age at accelerometer assessment, sex, education, ethnicity, Townsend deprivation index, BMI category, smoking status, alcohol drinking status, physical activity, diabetes, high blood pressure, depression, angina, stroke, heart attack, and depression.

*Table S 7 Hazard ratios of incident dementia by accelerometer-measured weekend catch-up sleep when using ≤0 hour as the reference group*

| Variable | N | HR (95%CI) | *P*-value |
|---|---|---|---|
| **By duration** | | | |
| ≤0 hour | 29413 | 1.00 (Reference) | |
| >0-0.5 hour | 15107 | 0.94 (0.77-1.14) | 0.523 |
| >0.5-1 hour | 13969 | 0.89 (0.72-1.10) | 0.271 |
| >1-1.5 hours | 10150 | **0.63 (0.47-0.84)** | **0.002** |
| >1.5-2 hours | 6458 | 0.82 (0.59-1.15) | 0.250 |
| >2 hours | 8679 | 0.82 (0.59-1.14) | 0.243 |
| **Binary** | | | |
| None or ≤0 hour | 29413 | 1.00 (Reference) | |
| >0 hour | 54363 | **0.85 (0.73-0.98)** | **0.029** |

The sensitivity analyses were based on Cox model adjusted for age at accelerometer assessment, sex, education, ethnicity, Townsend deprivation index, BMI category, smoking status, alcohol drinking status, physical activity, diabetes, high blood pressure, depression, angina, stroke, heart attack, and depression

*Figure S 1 Associations of accelerometer-measured weekend catch-up sleep with risk of incident dementia modelled using restricted cubic spline model*

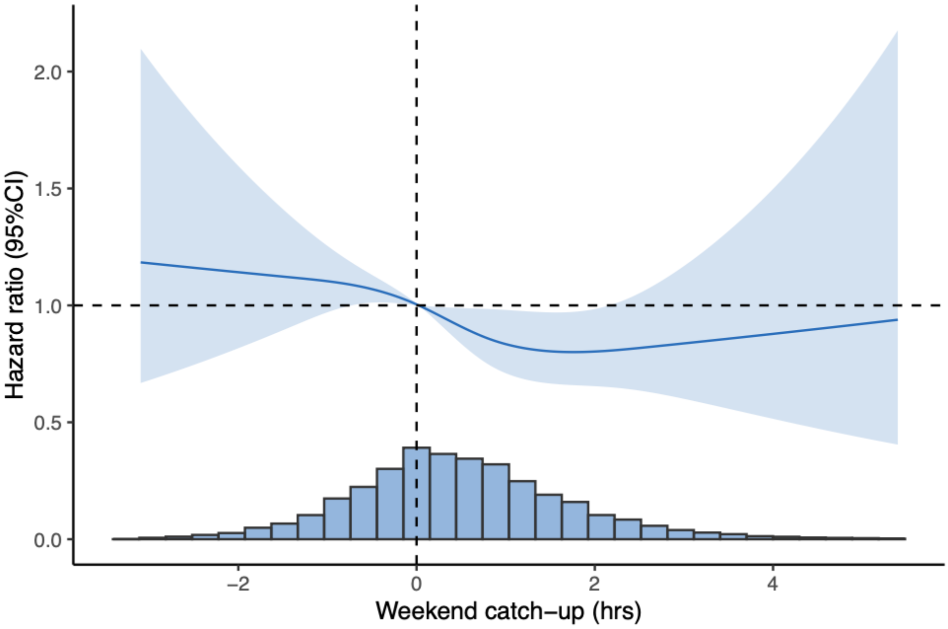

The Cox model was adjusted for age at accelerometer assessment, sex, education, ethnicity, Townsend deprivation index, BMI category, smoking status, alcohol drinking status, physical activity, sleep duration on weekday, shift work status, chronotype, diabetes, high blood pressure, depression, angina, stroke, heart attack, and depression. Reference point was set to be 0 hour on x-axis. Number of knots was chosen based on AIC criteria. *P* for linear trend=0.033, *P* for non-linear trend=0.563.

*Figure S 2 Hazard ratios of incident dementia by accelerometer-measured average sleep duration (A) and sleep durations on weekday (B) and weekend (C), modelled using restricted cubic spline model*

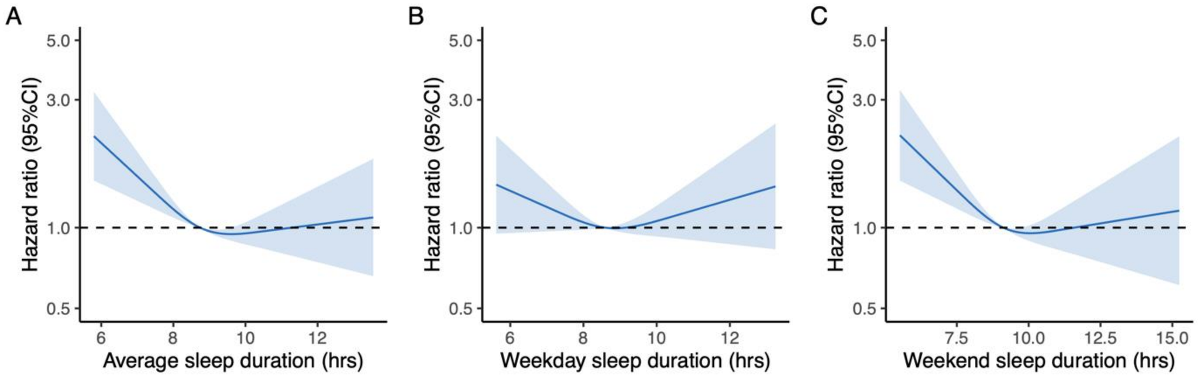

The Cox model was adjusted for age at accelerometer assessment, sex, education, ethnicity, Townsend deprivation index, BMI category, smoking status, alcohol drinking status, physical activity, diabetes, high blood pressure, depression, angina, stroke, heart attack, and depression. Reference point was set to be population median sleep duration on x-axis. Number of knots was chosen based on AIC criteria.

*Figure S 3 Hazard ratios of incident dementia accelerometer-measured weekend catch-up sleep (>0.5 hour versus less or none) in study subgroups*

| Subgroup | Case/PY | HR | P-interaction |
|---|---|---|---|
| Age | | | 0.466 |
| <65 | 83/368875.4 | 0.95 (0.62, 1.54) | |
| ≥65 | 630/299052.7 | 0.80 (0.68, 1.18) | |
| Sex | | | 0.532 |
| Women | 334/378498.2 | 0.86 (0.68, 1.26) | |
| Men | 379/289429.9 | 0.78 (0.63, 1.24) | |
| Social deprivation | | | 0.713 |
| High | 352/333070.4 | 0.84 (0.67, 1.25) | |
| Low | 361/334857.7 | 0.79 (0.64, 1.25) | |
| BMI | | | 0.116 |
| Normal/Underweight | 266/260654.6 | 0.95 (0.74, 1.28) | |
| Overweight/Obesity | 447/407273.5 | 0.74 (0.61, 1.22) | |
| Smoking | | | 0.426 |
| Ever smoking | 378/288965.0 | 0.77 (0.62, 1.24) | |
| Never smoking | 335/378963.1 | 0.87 (0.69, 1.25) | |

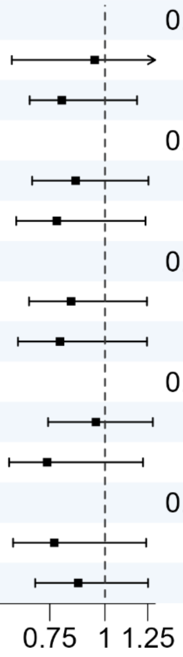

The Cox model was adjusted for age at accelerometer assessment, sex, education, ethnicity, Townsend deprivation index, BMI category, smoking status, alcohol drinking status, physical activity, diabetes, high blood pressure, depression, angina, stroke, heart attack, and depression. The stratification variables were not adjusted for the corresponding analyses. *P*-interactions were calculated from the likelihood ratio test comparing the model with and without the interaction term crossing weekend catch-up sleep and the stratification variable.
